# Laboratory Tests for Active Pulmonary Tuberculosis Diagnosis in Adults From Lower-Middle-Income Countries in Southeast Asia: A Systematic Review

**DOI:** 10.1101/2025.10.04.25337313

**Authors:** Josephine Abrazaldo, Patrick de Vera, Ferdinand A. Mortel

## Abstract

Pulmonary Tuberculosis remains a global public health problem in lower middle income countries of the ASEAN, where due to inadequate resources diagnostic limitations contribute to delayed detection and treatment. This systematic review aimed to evaluate the diagnostic accuracy of selected laboratory test used by these lower middle income countries. A structured literature search was conducted that assessed the sensitivity, specificity and predicted values of the selected diagnostic methods, adhering to the PRISMA guidelines and quality appraisal was done using QUADAS-2. Preliminary synthesis indicates that GeneXpert demonstrated high sensitivity and specificity as compared to conventional microscopy. TB LAMP present a feasible alternative in low resource countries.

## INTRODUCTION

Tuberculosis (TB) remains a global public health problem, affecting millions of individuals per year, especially in countries located in Africa, Southeast Asia, and the Western Pacific. ^1,2^ This infectious disease is also one of the leading causes of infection-related mortality, responsible for 1.3 million deaths in 2022 alone. ^2,3^ To end the global epidemic, the World Health Organization (WHO) identified priority countries for targeted intervention and support.

Most lower-middle-income countries of the Association of Southeast Asian Nations (ASEAN) were included in the TB high-burden list released by the WHO. ^1^ These countries are considered priority areas due to the high number of newly diagnosed TB cases yearly. Moreover, these countries are faced with the increasing burden of human immunodeficiency virus (HIV) co-infection and multi-drug resistant TB (MDR-TB). ^1^

Early diagnosis of TB is, therefore, essential to ensure timely intervention and prevent further disease transmission. While clinical diagnosis of TB is still relevant for early intervention, particularly in low-resource settings, its varying accuracy limits its utility.^16, 17^ Bacteriologic confirmation is still warranted to ensure correct treatment, especially due to the increasing trend in MDR-TB. ^2^ In recent years, more accurate and rapid diagnostic tests have been utilized in developed countries to improve patient prognosis and minimize disease transmission. ^4^

Nonetheless, countries with high TB burdens often have inadequate resources; thus, access to accurate diagnostic tests is still a problem. Traditional methods such as sputum smear microscopy and culture are still the commonly performed tests in least-developed countries. ^4^ Newer diagnostic tests had been developed; however, validation was limited to single settings and certain countries and regions. Moreover, the accuracy of these diagnostic tools is often affected by the prevalence of TB and co-infection with HIV, which is increasing nowadays. ^4,5^ This study, therefore, aims to determine the diagnostic tests for bacteriologic confirmation of TB, which were validated in low-resource settings, focusing on the ASEAN lower-middle-income countries. Specifically, this systematic review intends to synthesize the accuracy measures of these tests in diagnosing active pulmonary TB (PTB) among adults from Cambodia, Indonesia, Laos, Myanmar, Philippines, and Vietnam.

## METHODOLOGY

### Study design

The researchers conducted a systematic review for the qualitative synthesis of relevant studies using Preferred Reporting Items for Systematic reviews and Meta-Analyses (PRISMA) guidelines. The study was conducted from May 1 to June 30, 2025. This protocol was registered with the PROSPERO International prospective register of systematic reviews database (registration number CRD4 2025 1022894) on March 21,2025.

### Study population

The current study applied the following inclusion criteria: 1) study population includes adult patients who underwent any laboratory test for active PTB diagnosis using sputum samples only, 2) diagnostic accuracy studies which presented any the following parameters: discriminative ability, sensitivity, specificity, positive and negative predictive values, 3) study performed in any of the following ASEAN lower-middle-income countries: Cambodia, Indonesia, Laos, Myanmar, the Philippines and Vietnam, and 4) available full-text article.

Excluded articles were 1) studies focused on the diagnosis of extra pulmonary TB, latent TB, drug-resistant TB, 2) studies which utilize other respiratory samples e.g. pleural fluid and BAL samples 3) Laboratory test combined with clinical or immunologic markers, 4) results combined for adults and children, with no subgroup analysis, 5) case reports, case series, editorials and reviews, 6) systematic review and/or meta-analysis, 7) conference proceedings or abstracts with no access to full-text articles, and 8) articles with no available English translation or cannot be translated to English.

### Information sources and search strategy

Two independent researchers conducted a literature search of relevant studies from electronic databases including PubMed, LILACS, HERDIN, Google Scholar (first 300 articles), and ResearchGate (first 100 articles). Furthermore, hand-searching was performed to identify other articles. This started from April 12, 2025 to May 15, 2025. Corresponding authors was contacted for studies with no available full-text articles, including oral and poster presentations in symposia and conferences.

The following keywords and MESH terms were used for the search and Boolean operators were applied for an expanded search*: “tuberculosis”, “pulmonary tuberculosis”, “PTB”, “mycobacterium tuberculosis”, “sputum”, “diagnostic accuracy”, “diagnostic value”, “accuracy”, “sensitivity”, “specificity”, “predictive value”, “discriminative ability”, “c-statistics”.* No restriction with regard to the date of publication will be applied.

### Selection of Studies

Duplicate studies was identified using Mendeley Desktop App by Elsevier and were excluded from the review. Selected titles and abstracts were screened for eligibility by the two independent researchers. Disagreements were resolved by third reviewer. The two reviewers downloaded and reviewed the full-text versions of the selected articles to check for eligibility.

### Data Extraction and Management

The two independent reviewers performed the data extraction process. The following information was obtained from each study: authors, year of publication, country, study design (retrospective/prospective), sample size, case finding (active/ passive), sputum induction (%yes), sputum processing (%yes), index test/s (sputum smear microscopy/ specific molecular testing, reference standard used (culture/others), and outcomes. Demographic and clinical characteristics of patients include mean age, sex (% male), HIV status (%positive), previous history of TB (%yes), previous TB treatment (%yes), For all studies that were included in this systematic review culture is considered the reference method.

### Outcome variables and definition

#### Primary outcome

Sensitivity and specificity of the index tests in diagnosing active PTB

#### Secondary outcomes

1. Positive predictive value (PPV)
2. Negative predictive value (NPV)

### Assessment of risk of bias

An experienced epidemiologist used the QUADAS-2 tool of the Review Manager version 5.4 software (Appendix A) to appraise the risk of bias of each eligible study. Using this tool, each study was assessed in terms of the representativeness of samples, selection criteria, reference standard, and timing of outcome confirmation.

### Ethical considerations

Since the study used data from the results of previously published studies and no patient contact was performed, a certificate of exemption was issued by the MCU ERB.

## SEARCH RESULT

A total of 5821 journals were identified during the initial search, 5,393 from PubMed,3 from Herdin,25 from LILACS,300 from Google scholar and 100 from research gate as shown in Figure 1. Duplicate records removed before screening was 238. A total of 5583 journals were screened, 36 were sought for retrieval. 5 had no full-text available;thus, 31 full text were screened. 9 articles were found to be eligible based on the inclusion and exclusion criteria.

**Figure 1.**
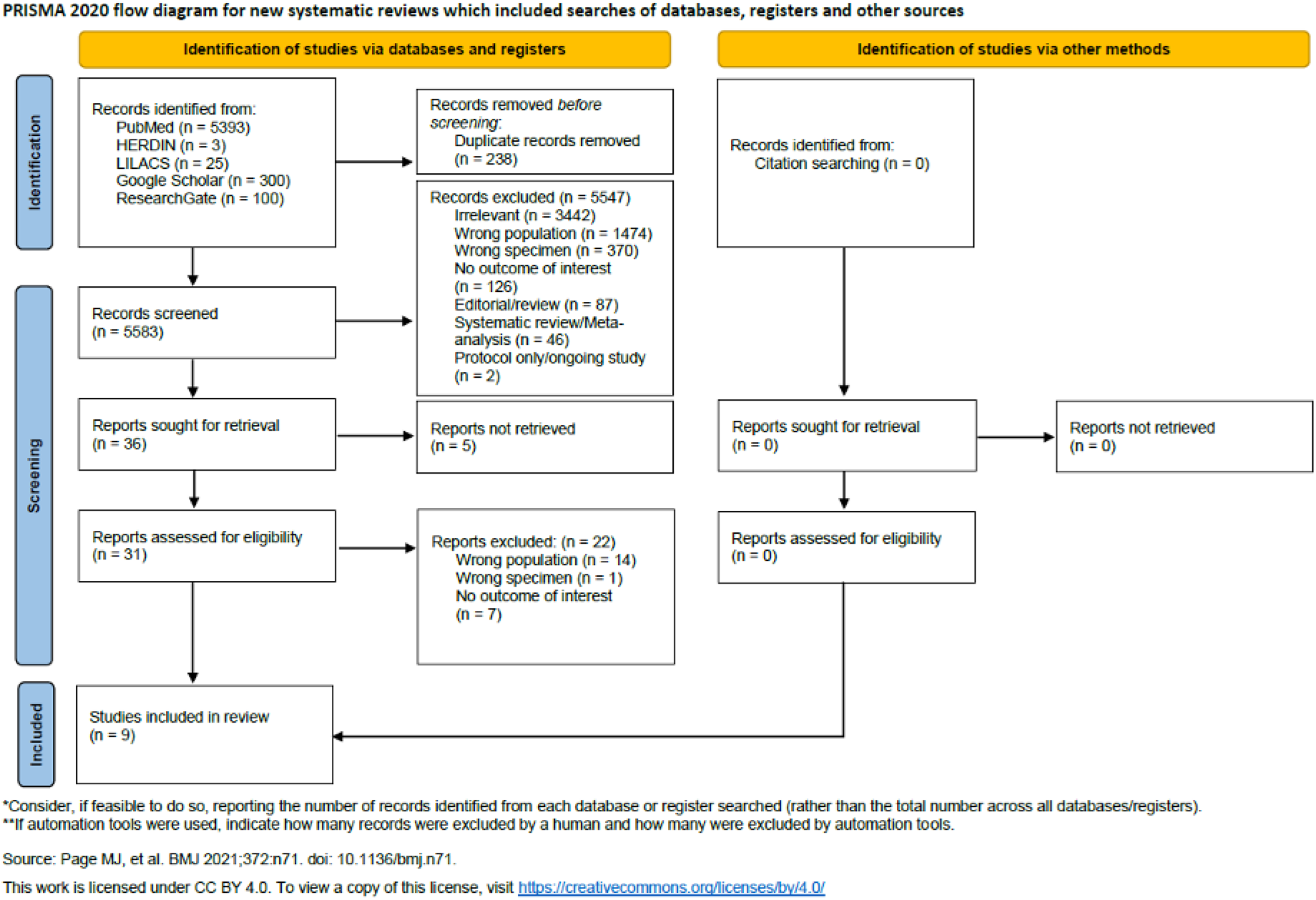
PRISMA Workflow

The included studies were published from 2003 - 2023, as detailed in Figure 1 Characteristics of the included studies are as follows; about half of the studies were conducted in Vietnam, ^6–9^ with two each from Indonesia^10,11^ and the Philippines^12,13^ and one from Cambodia^14^. Three studies were conducted across multiple countries;^6,7,12^ however subgroup analysis of each country were available hence were still included. All included studies employed a prospective study design. A total sample size across studies was 3,723 with individual sample sizes ranging from 100 to 1,179 participants.

Only one study reported the use of sputum induction, and it was performed only when deemed necessary.^11^ Sputum analyses were primarily conducted for culture based diagnostics using the NALC-NaOH decontamination method. Five studies evaluated the diagnostic accuracy of smear microscopy(SSM), four assessed GeneXpert, and three investigated TB LAMP. The study by Bellen 2023 focused on rapid diagnostic test - PhageTek Assay alongside SSM.^13^ Culture were mostly done using Lowenstein-Jensen (LJ) medium and Mycobacteria growth Indicator Tube (MGIT) systems.

Table 1 presents the characteristics of the included patients. The mean age ranged from 36.0 to 48.6 years across studies, but not all studies provided the median age. Between 50% and 70 % of participants were male. Among those with known HIV status, positivity ranged from 2% to 47%. A history of tuberculosis or prior treatment was reported in 0% to 64% of patients.

**Table 1.** Characteristics of included studies.

| Author/year | Country | Study design | Sample size | Case finding | Sputum induction | Sputum processing | Index test/s | Reference standard | Outcomes |
| --- | --- | --- | --- | --- | --- | --- | --- | --- | --- |
| Karuniawati 2023 | Indonesia | Prospective | 447 | Passive | Yes, if necessary | *Culture: NALC-NaOH | SSM, GeneXpert | Culture (LJ/ Ogawa/ MGIT) | Sensitivity, Specificity |
| Cheng 2020 | Cambodia | Prospective | 499 | Passive | NR | *SSM/ TB-LAMP: none<br>*Culture/ GeneXpert: NALC-NaOH | SSM, GeneXpert, TB-LAMP | Culture (LJ/ MGIT) | Sensitivity, Specificity |
| Wahid 2020 | Indonesia | Prospective | 100 | Passive | No | Culture: Petroff method | TB-LAMP | Culture (LJ) | Sensitivity, Specificity, PPV, NPV |
| Pham 2018 | Vietnam<sup>a</sup> | Prospective | 299<sup>b</sup> | Passive | NR | *Culture/ GeneXpert: NALC-NaOH | GeneXpert, TB-LAMP | Culture (LJ/ MGIT) | Sensitivity, Specificity |
| Nabeta, 2017 | Vietnam<sup>a</sup> | Prospective | 270<sup>b</sup> | Passive | NR | *SSM/Culture : NALC-NaOH | SSM | Culture (LJ/ MGIT) | Sensitivity |
| Boehme 2011 | Philippines<sup>a</sup> | Prospective | 387<sup>b</sup> | Passive | NR | Culture: NALCNaOH, Petroff method | GeneXpert | Culture (LJ/ Ogawa) | Sensitivity, Specificity, PPV, NPV |
| Maloney 2006 | Vietnam | Prospective | 1179 | Active | NR | Culture: oxalic acid | SSM | Culture (BACTEC 12B/ LJ) | Sensitivity, Specificity, PPV, NPV |
| Laserson 2006 | Vietnam | Prospective | 338 | Active | NR | *SSM: CB-18<br>*Culture: NaOH-NALC, oxalic acid | SSM | Culture (BACTEC 12B/ LJ) | Sensitivity, Specificity, PPV, NPV |
| Bellen 2003 | Philippines | Prospective | 204 | Passive | NR | SSM/PhageTek assay/Culture : NALC-NaOH | SSM, PhageTek assay (rapid test) | Culture (LJ) | Sensitivity, Specificity, PPV, NPV |

NPV: Negative Predictive Value; NR: Not reported; PPV: Positive Predictive Value; SSM: Sputum smear microscopy
<sup>a</sup>Research was conducted in multiple countries, but only data of target country was included <sup>b</sup>Only total number of patients from target country included in the analysis of accuracy was recorded in this table

**Table 2.** Characteristics of patients in included studies.

| Author/year | Mean age | Sex (%male) | HIV status %positive | Previous history of TB (%yes) | Previous TB treatment (%yes) |
| --- | --- | --- | --- | --- | --- |
| Karuniawati 2023 | 41.3 | 61% | NR | 42% | 42% |
| Cheng 2020 | NR<br>*Age groups reported instead. Most (24.2%) were ≥65 years | 56% | 11%<br>*3 out of 28 with known HIV status | NR | 27% |
| Wahid 2020 | NR<br>*Age groups reported instead. Most (49%) were 45-64 years | 65% | 7%<br>*2 out of 29 with known HIV status | 0% | 0% |
| Pham 2018 | 43.3* | 70% | 2% | NR | NR |
| Nabeta 2017 | 36<sup>a</sup> | 63%<sup>a</sup> | NR | NR | NR |
| Bohme 2011 | 46.3*<sup>a</sup> | 64%<sup>a</sup> | 47%<br>*8 out of 17 with known HIV status | NR | NR |
| Maloney 2006 | 48.6 | 50% | NR | 15% | 15% |
| Laserson 2005 | NR | NR | 6% | NR | NR |
| Bellen 2003 | 41.2 | 67% | NR | NR | 64% |

NR: Not reported; \*Median converted to mean using method by Wan et al ([MB1])<sup>a</sup> Characteristics of all enrolled patients, not just target population included in the analysis of accuracy

### Over all accuracy of laboratory tests in detecting active pulmonary tuberculosis

Six studies involving 2,937 participants assessed SSM’s performance in detecting PTB. Sensitivity varied widely, ranging from 34.4% to 89.3% and specificity ranging from 60.4% to 98.2%. The highest sensitivities were reported in Bellen 2003 (Philippines) and Karuniawati 2023 (Indonesia),^11,13^ while the remaining studies three from Vietnam, one from Cambodia, and two focused on U.S.-bound immigrants showed < 70% sensitivity, with Maloney 2006 having reported the lowest sensitivity 34.4%.^9^ Although specimen processing modestly enhanced sensitivity, overall results remained sub optimal. Specificity of SSM was generally high > 85% though it fluctuated depending on the staining technique. PPV and NPV varied considerably as it is influenced by disease prevalence and if population were actively screened or symptomatically tested. Factors such as smear processing (direct vs concentrated), type of stain use (auramine vs Ziehl-neelsen) had noticeable but inconsistent effects on diagnostic yield.

GeneXpert demonstrated consistently high diagnostic accuracy for pulmonary tuberculosis across the reviewed studies. Sensitivity ranged from 77.7% to 97.4%, with the highest values observed in studies by Karuniawati 2023, Cheng 2020, and Boehme 2011, indicating its strong capability to detect true TB cases.^11,12,14^ Specificity also remained high in most cases, particularly in Cheng and Boehme’s studies (96.8% and 97.9%, respectively). ^12,14^ However, it declined in specific study sites, including those documented by Pham 2018 (73.1%) and Karuniawati 2023 (73.3%), possibly due to number of specimen submitted for Karuniawati 2023 make used of pooled sputum while Pham 2 sputum sample.^6,11^ Notably, Boehme 2011 provided the most complete performance profile, reporting both positive predictive value (95.7%) and negative predictive value (95.9%),^12^ underscoring GeneXpert’s reliability for both confirming and ruling out TB.

TB LAMP across studies shows high specificity with Cheng 2020 and Pham showing the highest 97.4% and 96.9% respectively suggesting high potential in confirming TB cases in resource limited settings.^14^ Sensitivity exhibits considerable variations (71-100%) which may be due to sample preparation as in the case of Cheng morning specimen was used while Wahid 2020 used mixed specimen. ^6,10,14^

The diagnostic performance of the PhageTek rapid test, as reported by Bellen 2003, illustrates a low sensitivity and high specificity based on the threshold used for interpretation.^13^ A cutoff of ≥20 plaques is applied as defined by the manufacturer, the test achieves high specificity (86.1%) but low sensitivity (31.1%), suggesting it is more effective at confirming TB cases than detecting them, thus minimizing false positives but potentially missing many true cases. In contrast, lowering the threshold to detect “any plaque” increases sensitivity to 70.9%, enhancing its capacity to identify TB infections, but at the expense of specificity, which drops to 61.4%. This implies a higher rate of false positives, which could lead to unnecessary follow-up testing or treatment. Table 3 presents the summary of accuracy measures of the SSM, GeneXpert, TB - LAMP, and Phage Tek assay in detecting active PTB.

**Table 3.**
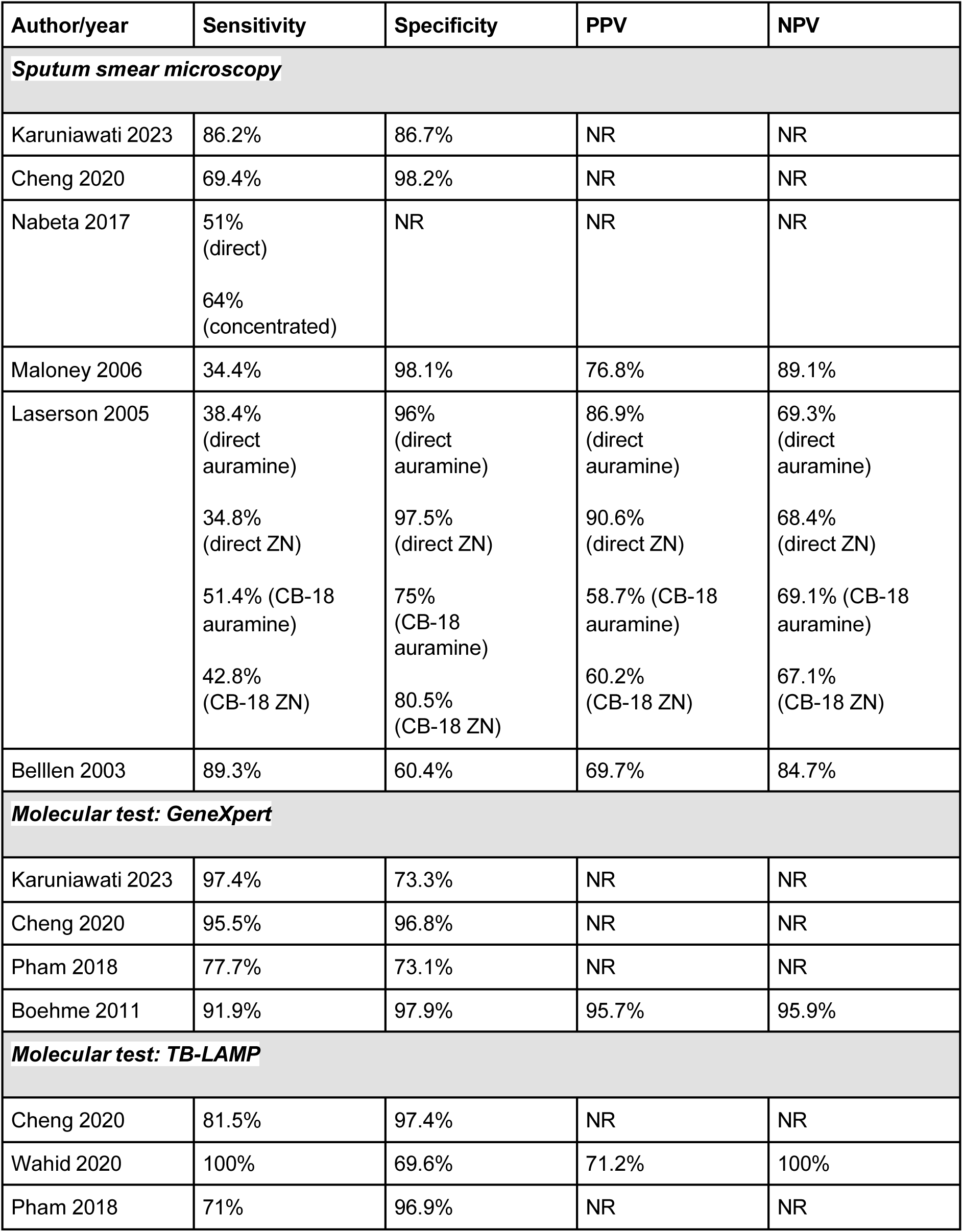
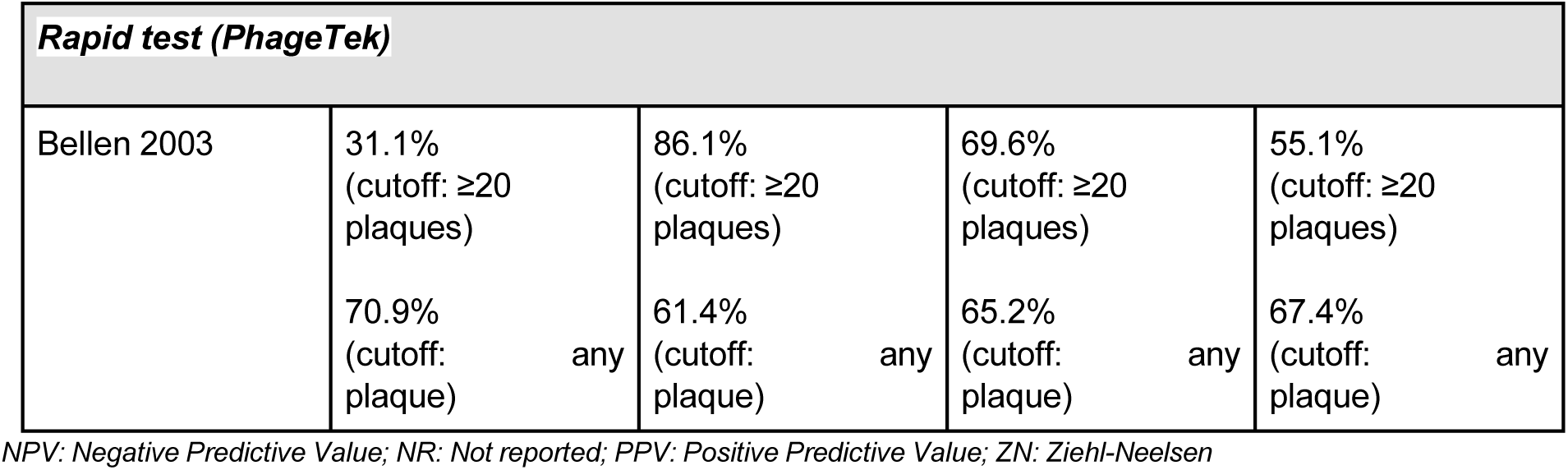
Accuracy of Laboratory tests in detecting active pulmonary tuberculosis: Overall.

## SUBGROUP ANALYSES

### Accuracy of Laboratory Test by Year of Publication

From 2003-2006 there is no clear improvement in the diagnostic accuracy for SSM across studies (table 4). But from 2011 to present (Table 5), there is a visible improvement in the SSM sensitivity from 51-64% in 2017 to 86.2% by 2023, possibly due to enhance staining protocols or laboratory practices. On the other hand, specificity remained high. GeneXpert remains high and show consistently high sensitivity 91.9-97.4% from 2011 onward though specificity slightly lower in the later studies which could be related to study population and specimen type. TB LAMP demonstrate variable sensitivity (71-100%) with generally high specificity except in Wahid 2020. ^10^. None of the included studies were published in 1999 or earlier.

**Table 4.** Accuracy of laboratory tests in detecting active pulmonary tuberculosis by year of publication (2000 to 2010)

| Author/year | Sensitivity | Specificity | Discriminative ability (AUC) | PPV | NPV |
| --- | --- | --- | --- | --- | --- |
| ***Sputum smear microscopy*** |  |  |  |  |  |
| Maloney 2006 | 34.4% | 98.1% | NR | 76.8% | 89.1% |
| Laserson 2005 | 38.4%<br>(direct auramine) | 96%<br>(direct auramine) | NR | 86.9%<br>(direct auramine) | 69.3%<br>(direct auramine) |
|  | 34.8%<br>(direct ZN) | 97.5%<br>(direct ZN) |  | 90.6%<br>(direct ZN) | 68.4%<br>(direct ZN) |
|  | 51.4% (CB-18 auramine) | 75%<br>(CB-18 auramine) |  | 58.7% (CB-18 auramine) | 69.1%<br>(CB-18 auramine) |
|  | 42.8%<br>(CB-18 ZN) | 80.5%<br>(CB-18 ZN) |  | 60.2%<br>(CB-18 ZN) | 67.1%<br>(CB-18 ZN) |
| Bellen 2003 | 89.3% | 60.4% | NR | 69.7% | 84.7% |
| ***Rapid test (PhageTek)*** |  |  |  |  |  |
| Bellen 2003 | 31.1%<br>(cutoff: ≥20 plaques) | 86.1%<br>(cutoff: ≥20 plaques) | NR | 69.6%<br>(cutoff: ≥20 plaques) | 55.1%<br>(cutoff: ≥20 plaques) |
|  | 70.9%<br>(cutoff: any plaque) | 61.4%<br>(cutoff: any plaque) |  | 65.2%<br>(cutoff: any plaque) | 67.4%<br>(cutoff: any plaque) |

**Table 5.** Accuracy of laboratory tests in detecting active pulmonary tuberculosis (2011 to present)

| Author/year | Sensitivity | Specificity | Discriminative ability (AUC) | PPV | NPV |
| --- | --- | --- | --- | --- | --- |
| ***Sputum smear microscopy*** |  |  |  |  |  |
| Karuniawati 2023 | 86.2% | 86.7% | NR | NR | NR |
| Cheng 2020 | 69.4% | 98.2% | NR | NR | NR |
| Nabeta 2017 | 51% (direct)<br>64% (concentrated) | NR | NR | NR | NR |
| ***Molecular test: GeneXpert*** |  |  |  |  |  |
| Karuniawati 2023 | 97.4% | 73.3% | NR | NR | NR |
| Cheng 2020 | 95.5% | 96.8% | NR | NR | NR |
| Pham 2018 | 77.7% | 73.1% | NR | NR | NR |
| Boehme 2011 | 91.9% | 97.9% | NR | 95.7% | 95.9% |
| ***Molecular test: TB-LAMP*** |  |  |  |  |  |
| Cheng 2020 | 81.5% | 97.4% | NR | NR | NR |
| Wahid 2020 | 100% | 69.6% | NR | 71.2% | 100% |
| Pham 2018 | 71% | 96.9% | NR | NR | NR |

### Accuracy of laboratory test in detecting active pulmonary tuberculosis by country

No study from Myanmar and Laos were included in this review.

### Cambodia

In Cambodia SSM showed high specificity, lower sensitivity. Its lower sensitivity affects early detection of PTB. GeneXpert on the other hand have high sensitivity and specificity which makes it an excellent choice for detecting PTB. TB LAMP strong sensitivity and specificity makes it a valuable tool for early detection of PTB for low resource settings country like Cambodia

**Table 6.** Accuracy of laboratory tests in detecting active pulmonary tuberculosis: Cambodia.

| Author/year | Sensitivity | Specificity | Discriminative ability (AUC) | PPV | NPV |
| --- | --- | --- | --- | --- | --- |
| ***Sputum smear microscopy*** |  |  |  |  |  |
| Cheng 2020 | 69.4% | 98.2% | NR | NR | NR |
| ***Molecular test: GeneXpert*** |  |  |  |  |  |
| Cheng 2020 | 95.5% | 96.8% | NR | NR | NR |
| ***Molecular test: TB-LAMP*** |  |  |  |  |  |
| Cheng 2020 | 81.5% | 97.4% | NR | NR | NR |

### Indonesia

Comparative diagnostic accuracy of PTB test in Indonesia as shown in Table 7 suggest that GeneXpert and TB LAMP with very high sensitivity but moderate specificity can have high false positive that can affect treatment decisions. While SSM remains valuable with moderately high diagnostic accuracy.

**Table 7.** Accuracy of laboratory tests in detecting active pulmonary tuberculosis: Indonesia.

| Author/year | Sensitivity | Specificity | Discriminative ability (AUC) | PPV | NPV |
| --- | --- | --- | --- | --- | --- |
| ***Sputum smear microscopy*** |  |  |  |  |  |
| Karuniawati 2023 | 86.2% | 86.7% | NR | NR | NR |
| ***Molecular test: GeneXpert*** |  |  |  |  |  |
| Karuniawati 2023 | 97.4% | 73.3% | NR | NR | NR |
| ***Molecular test: TB-LAMP*** |  |  |  |  |  |
| Wahid 2020 | 100% | 69.6% | NR | 71.2% | 100% |

### Philippines

GeneXpert laboratory test used in detecting active PTB in the Philippines showed optimal sensitivity and specificity (Boehme 2011)_12,_ although study of Bellen(2003)_13_ shows lower specificity this may be due to differences in test conditions. Phage Tek rapid test lacks the reliability as a standalone diagnostic tool for PTB.

**Table 8.**
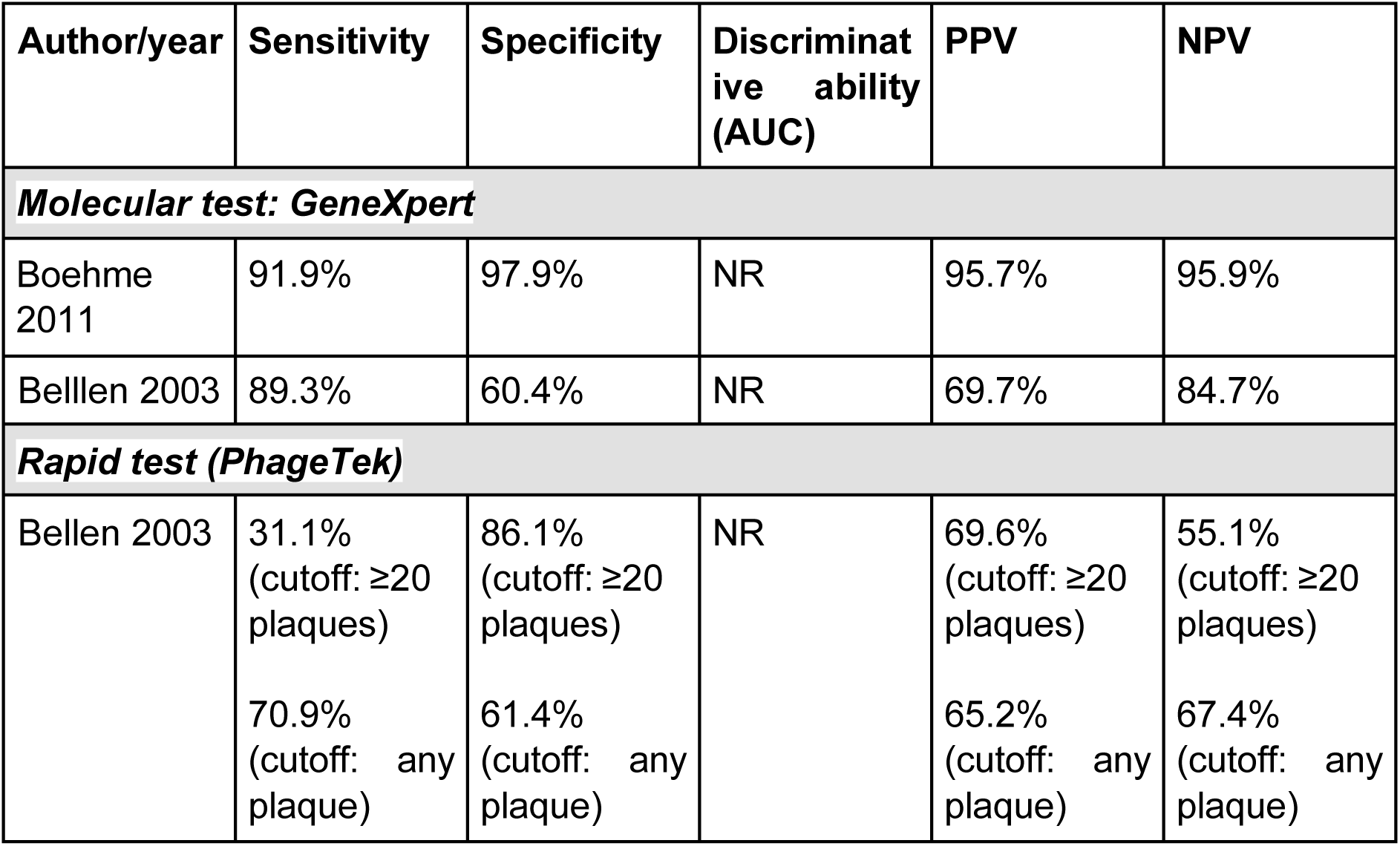
Accuracy of laboratory tests in detecting active pulmonary tuberculosis: Philippines.

### Vietnam

Sensitivity of SSM method range from 34-64% which means that it often miss true TB cases while specificity ranges from 75% to >97 % indicating its strong capability to rule out TB when negative. GeneXpert shows high sensitivity with moderate specificity while TB LAMP offers a high specificity that can be useful in confirmatory settings.

**Table 9.** Accuracy of laboratory tests in detecting active pulmonary tuberculosis: Vietnam.

| Author/year | Sensitivity | Specificity | Discriminative ability (AUC) | PPV | NPV |
| --- | --- | --- | --- | --- | --- |
| Sputum smear microscopy |  |  |  |  |  |
| Nabeta 2017 | 51% (direct)<br>64% (concentrated) | NR | NR | NR | NR |
| Maloney 2006 | 34.4% | 98.1% | NR | 76.8% | 89.1% |
| Laserson 2005 | 38.4% (direct auramine) | 96% (direct auramine) | NR | 86.9% (direct auramine) | 69.3% (direct auramine) |
|  | 34.8% (direct ZN) | 97.5% (direct ZN) |  | 90.6% (direct ZN) | 68.4% (direct ZN) |
|  | 51.4% (CB-18 auramine) | 75% (CB-18 auramine) |  | 58.7% (CB-18 auramine) | 69.1% (CB-18 auramine) |
|  | 42.8% (CB-18 ZN) | 80.5% (CB-18 ZN) |  | 60.2% (CB-18 ZN) | 67.1% (CB-18 ZN) |
| Molecular test: GeneXpert |  |  |  |  |  |
| Pham 2018 | 77.7% | 73.1% | NR | NR | NR |
| Molecular test: TB-LAMP |  |  |  |  |  |
| Pham 2018 | 71% | 96.9% | NR | NR | NR |

## RISK OF BIAS

Based on the QUADAS −2 risk of bias summary (Figure 2&3). Most studies demonstrate low risk of bias across all domains including patient selection, index test conduct, reference standard interpretation, and flow / timing. This suggest high internal validity and confidence in diagnostic performance outcomes. However Wahid (2020) have a high risk of bias and high applicability concern in the patient selection domain. This may reflect design limitations, Wahid et al excluded patients with suspected MDR-TB which may lead to selection bias.Transparency was also appraised using PRISMA 2020 checklist.

**Figure 2.**
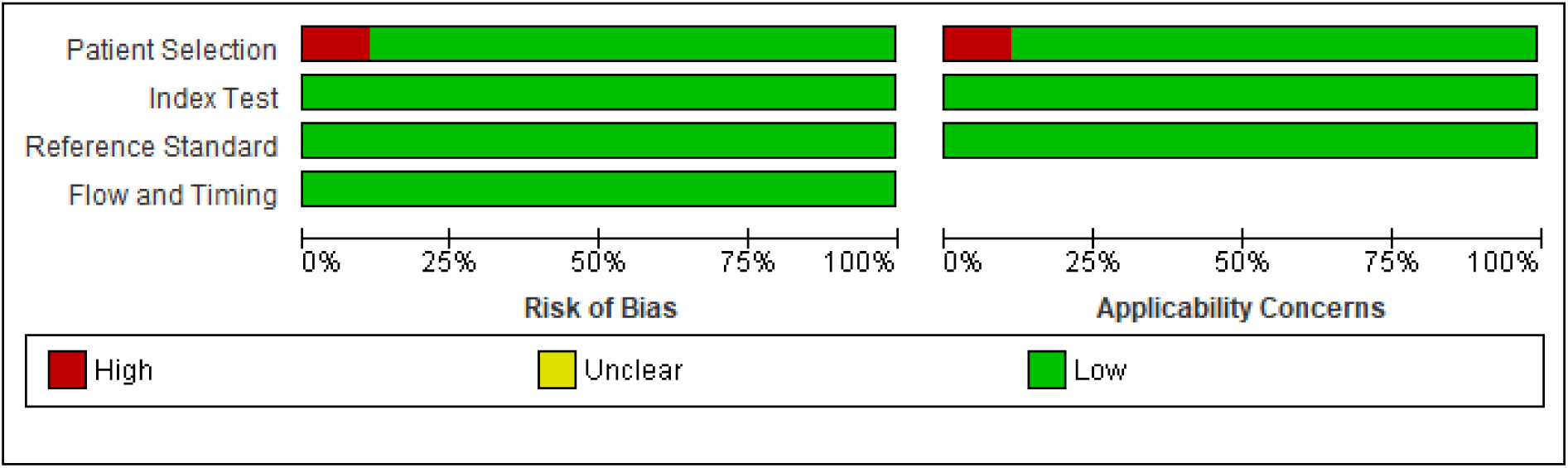
Risk of bias and applicability concerns graph: review authors’ judgements about each domain presented as percentages across included studies

**Figure 3.**
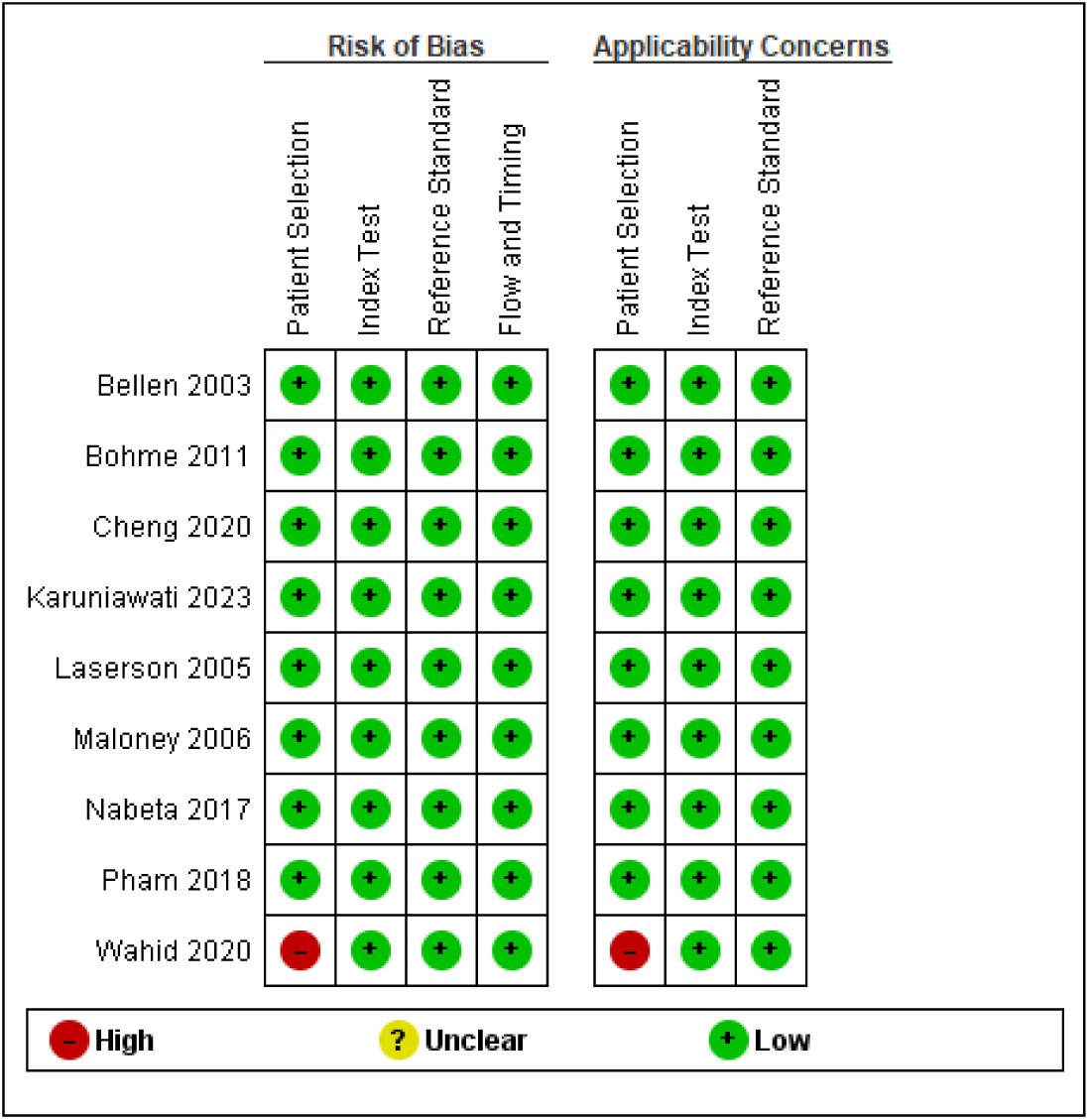
Risk of bias and applicability concerns summary: review authors’ judgements about each domain for each included study

## DISCUSSION

This detailed systematic review evaluated the diagnostic performance of SSM, GeneXpert and TB LAMP as well as the Phage Tek rapid test for detection of active PTB among adults, incorporating data from Vietnam,Cambodia, Indonesia and the Philippines. Results suggest that while each laboratory test serves a particular role, molecular test provide superior sensitivity and wider case detection than traditional microscopy, but in a low resource setting such as Vietnam, Cambodia, Indonesia and the Philippines TB LAMP can be ideal. Liping et al state that the low cost and accuracy of LAMP make it suitable for use in developing countries with high incidence of PTB.^15^ GeneXpert consistently demonstrates high sensitivity from 77.7% in Vietnam to 97.4% in Indonesia, confirming its capability in detecting true TB cases. Specificity varies from 60.4% and 97.9% in the Philippines and 73.1% in Vietnam. Predictive value is high with PPV of 95.7% and NPV 95.9%.^12^ This may be due to study population and prevalence rate. TB LAMP shows remarkable specificity value (up to 96.9%) in Vietnam and variable sensitivity from (71%) Vietnam to 100 %. The study of Wahid 2020 also reported a 100% NPV suggesting reliability in ruling out TB cases.^10^ SSM remains the most variable tool across regions with a sensitivity of 34.4 (Lowest in Vietnam) as stated in the study of Maloney 2006 ^9^ while Indonesia 86.2% and Philippines 69.4%. On the other hand specificity is high across all countries exceeding 95%. Phage Tek was only done in the Philippines and is dependent in its cut off as given by the manufacturer. The sensitivity range from 31.1% for plaques more than 20 and 70.9% for any plaques. Specificity ranging from 61.4 to 86.1%. PPV and NPV indicate limited diagnostic utility.

## Conclusion

Based on the data gathered across different settings and populations, Molecular diagnostic GeneXpert and TB LAMP outperform SSM in terms of sensitivity reinforcing their critical role in improving TB case detection. While GeneXpert consistently outperforms TB LAMP and SSM as diagnostic tool, TB LAMP can be an alternative assay in resource-limited countries due to its higher specificity making it highly reliable for ruling out TB. Sputum Smear Microscopy though accessible, cost-effective, and specific, demonstrates consistenty low sensitivity, making it inadequate as a standalone diagnostic tool in high TB burden countries. Furthermore its performance variability was observed across countries. The researchers recommends that 1) SSM can still be used in a low-resource setting however as there are many factors involved in the accurate diagnosis of Pulmonary Tuberculosis, Ensuring the proficiency of the Medical Technologist and standardization of sputum processing can help improve the diagnostic capability of SSM 2) TB LAMP can be a great substitute for Gene Xpert in terms of cost effectiveness as well as its high specificity..

## Data Availability

All data produced in the present study are available upon reasonable request to the authors
All data produced in the present work are contained in the manuscript

## APPENDIX A. Details of bias assessment

### Karuniawati 2023

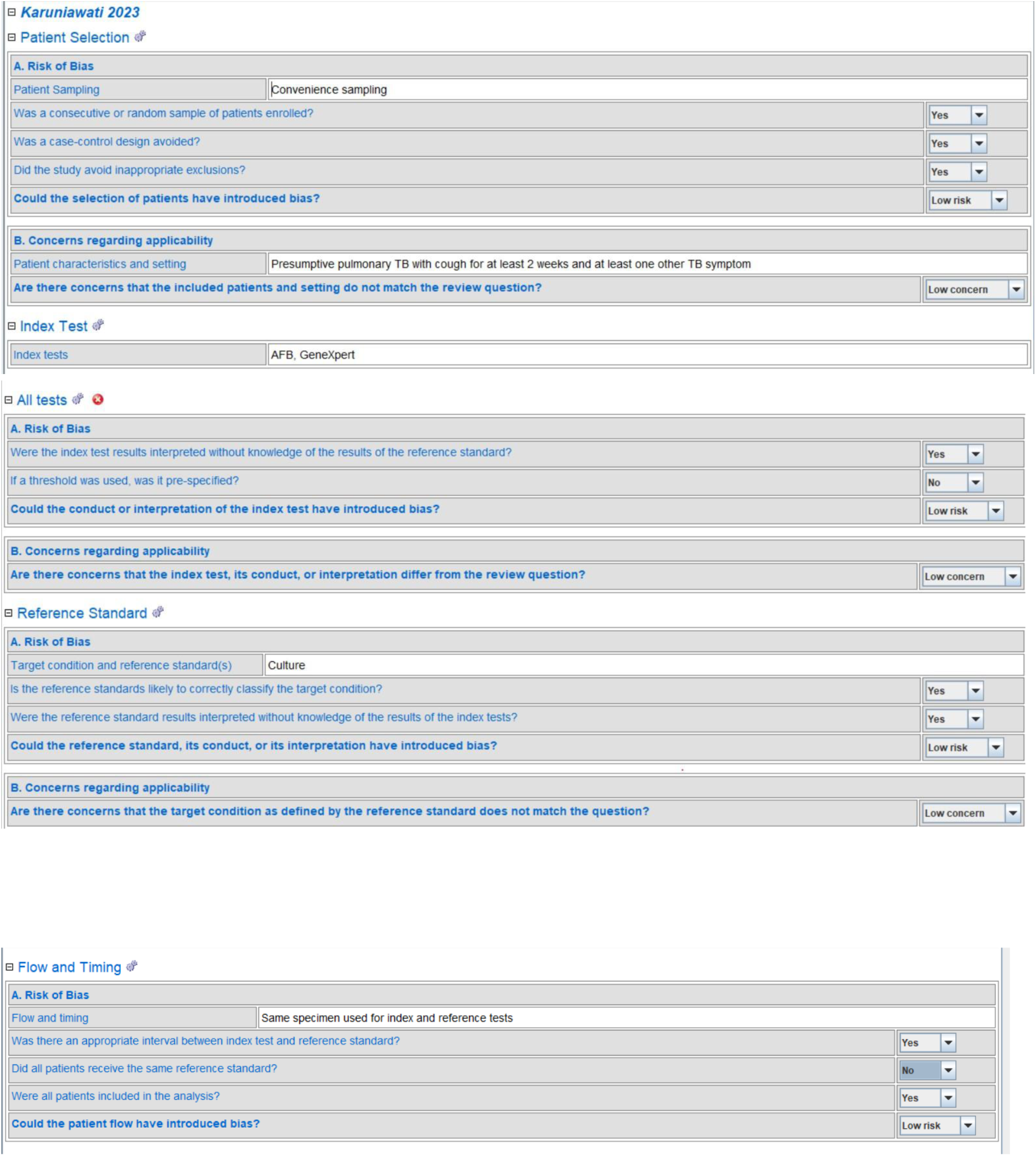

### Cheng 2020

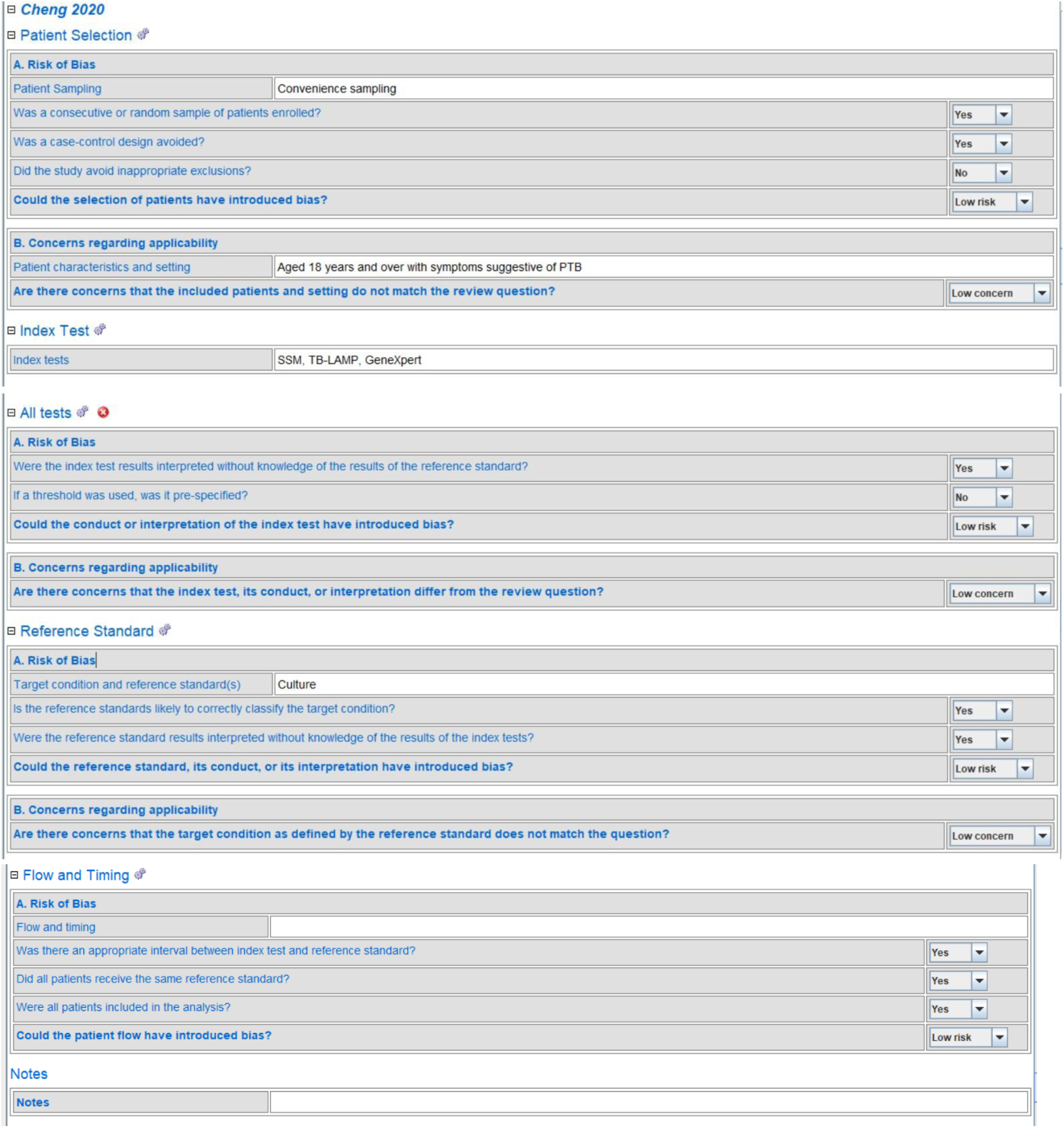

### Wahid 2020

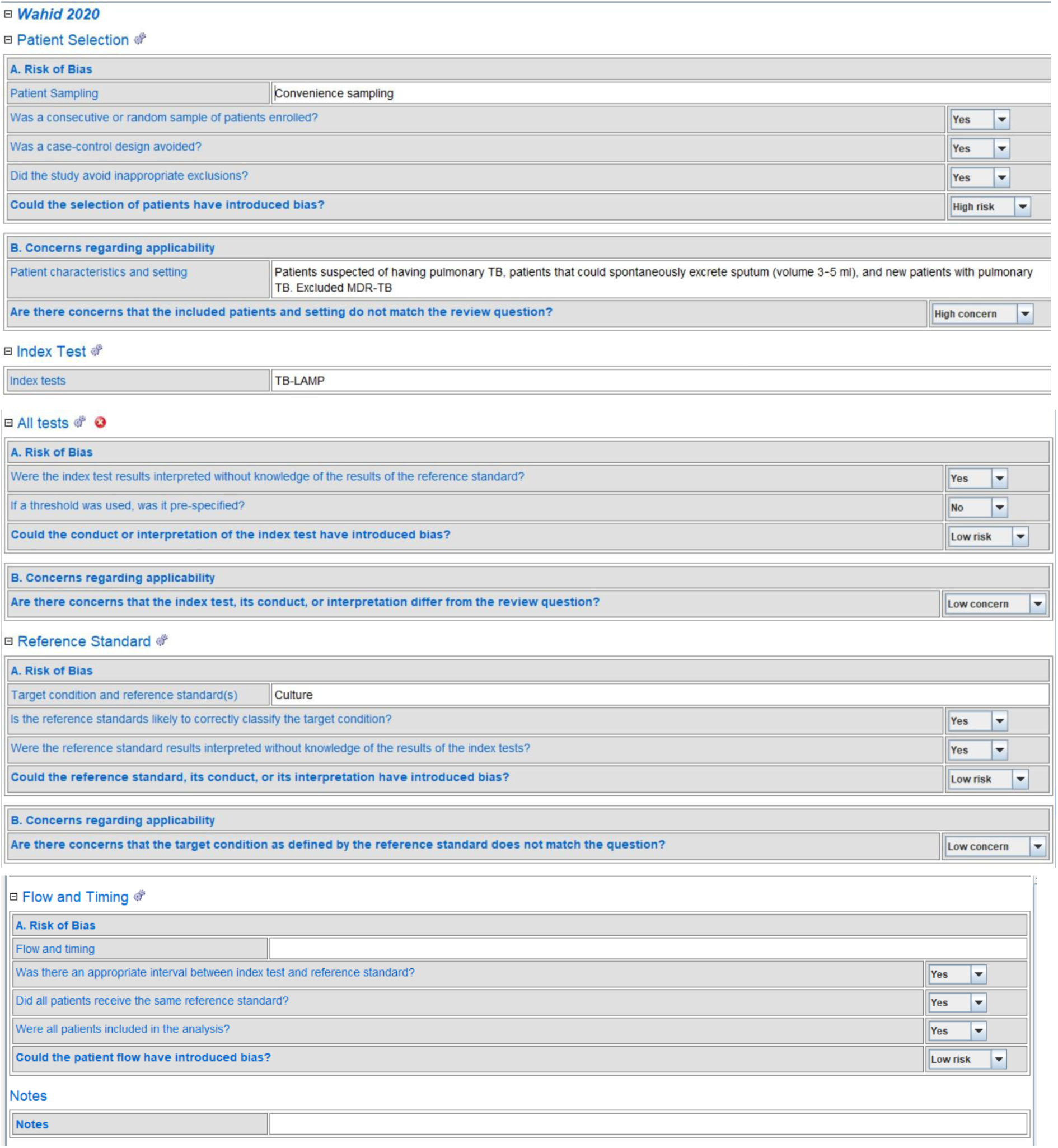

### Pham 2018

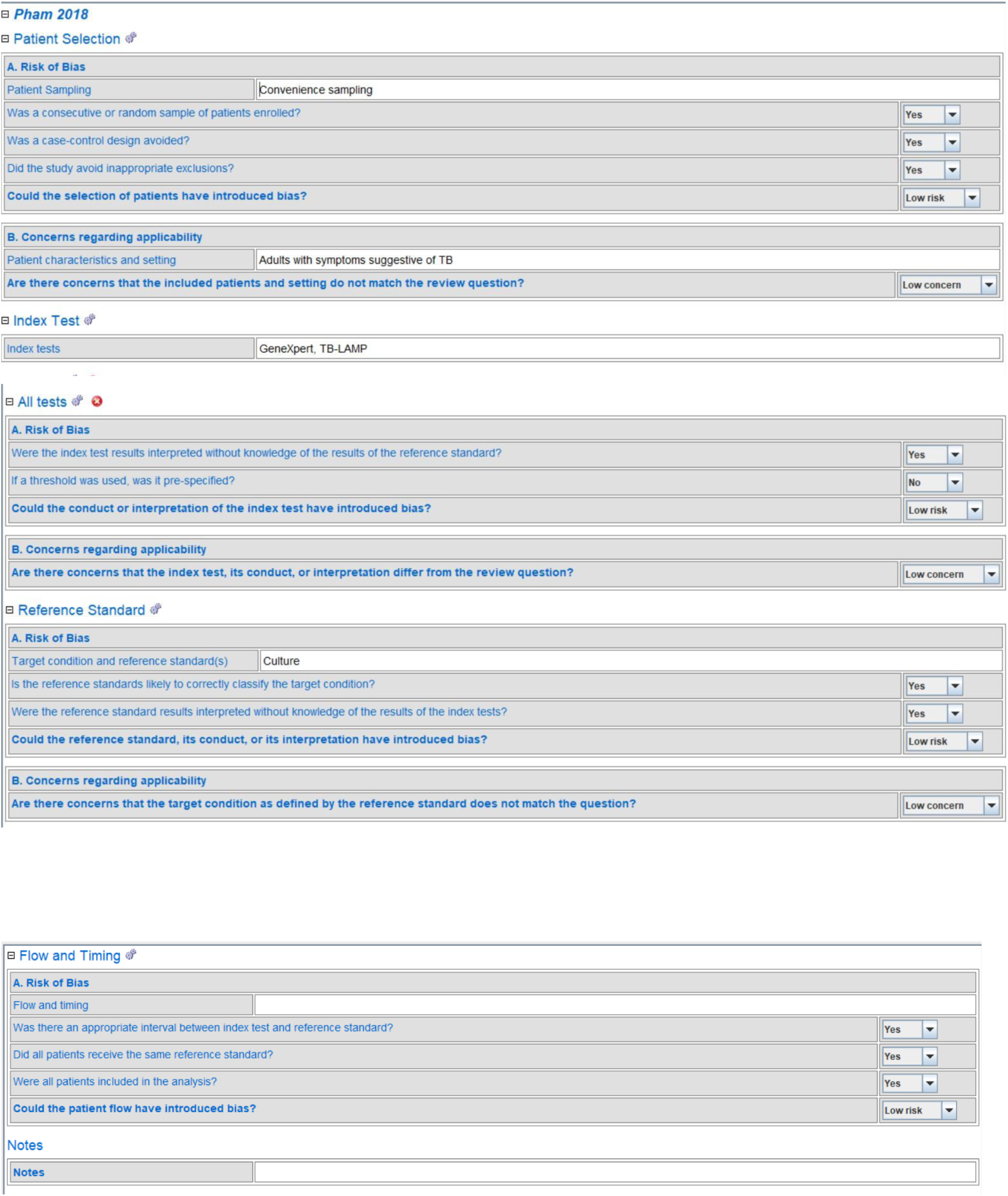

### Maloney 2006

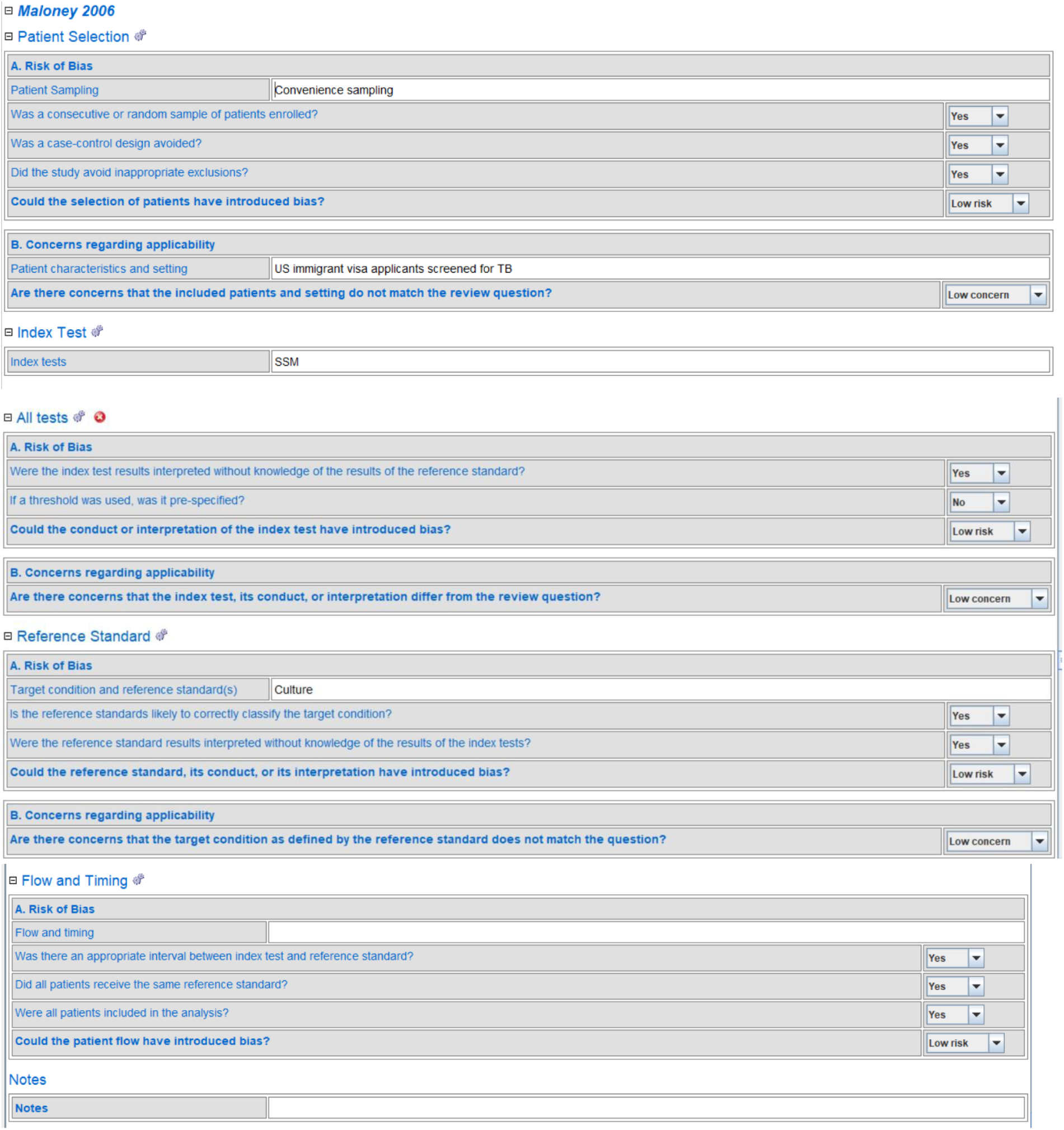

### Nabeta 2017

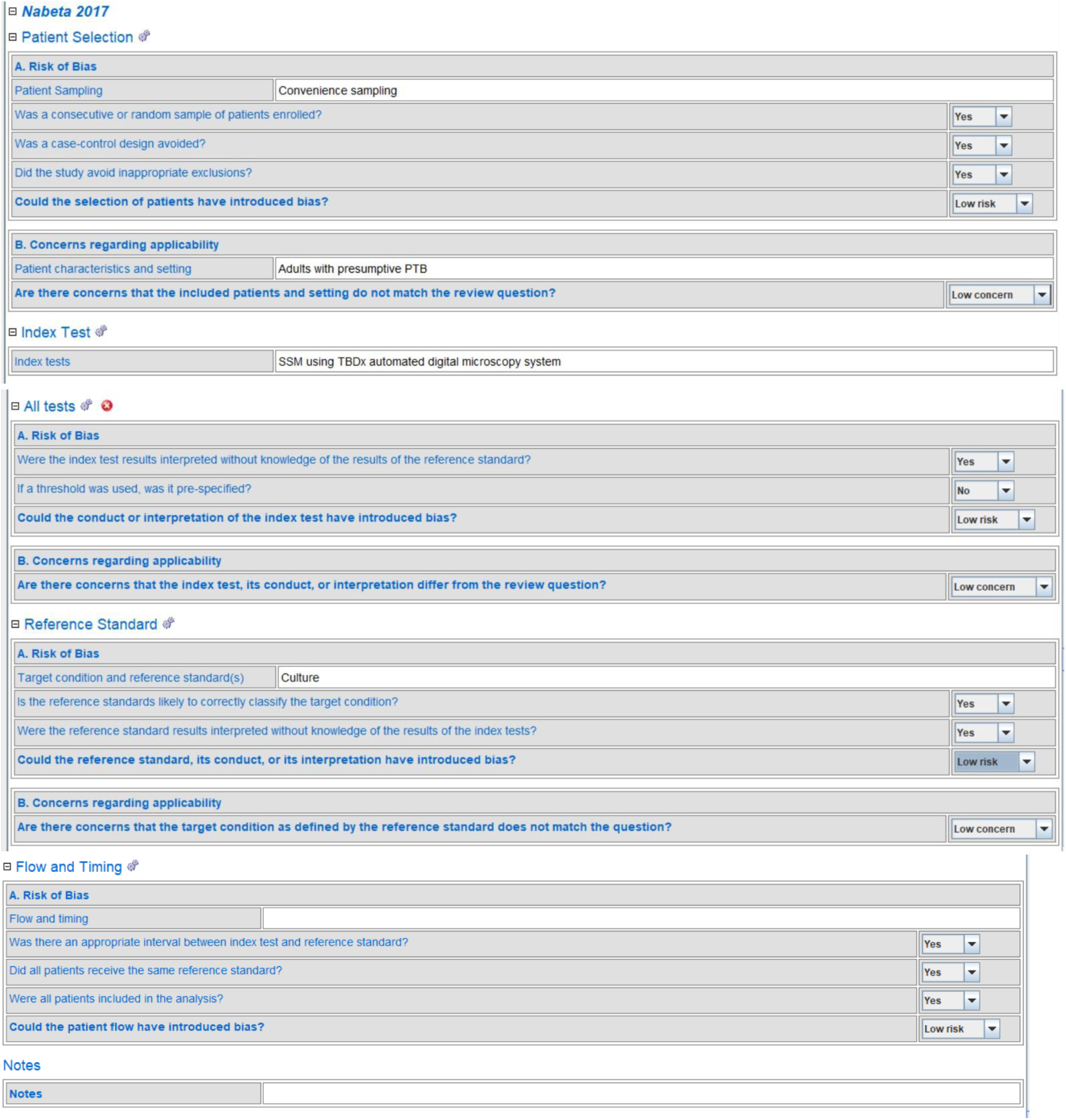

### Boehme 2011

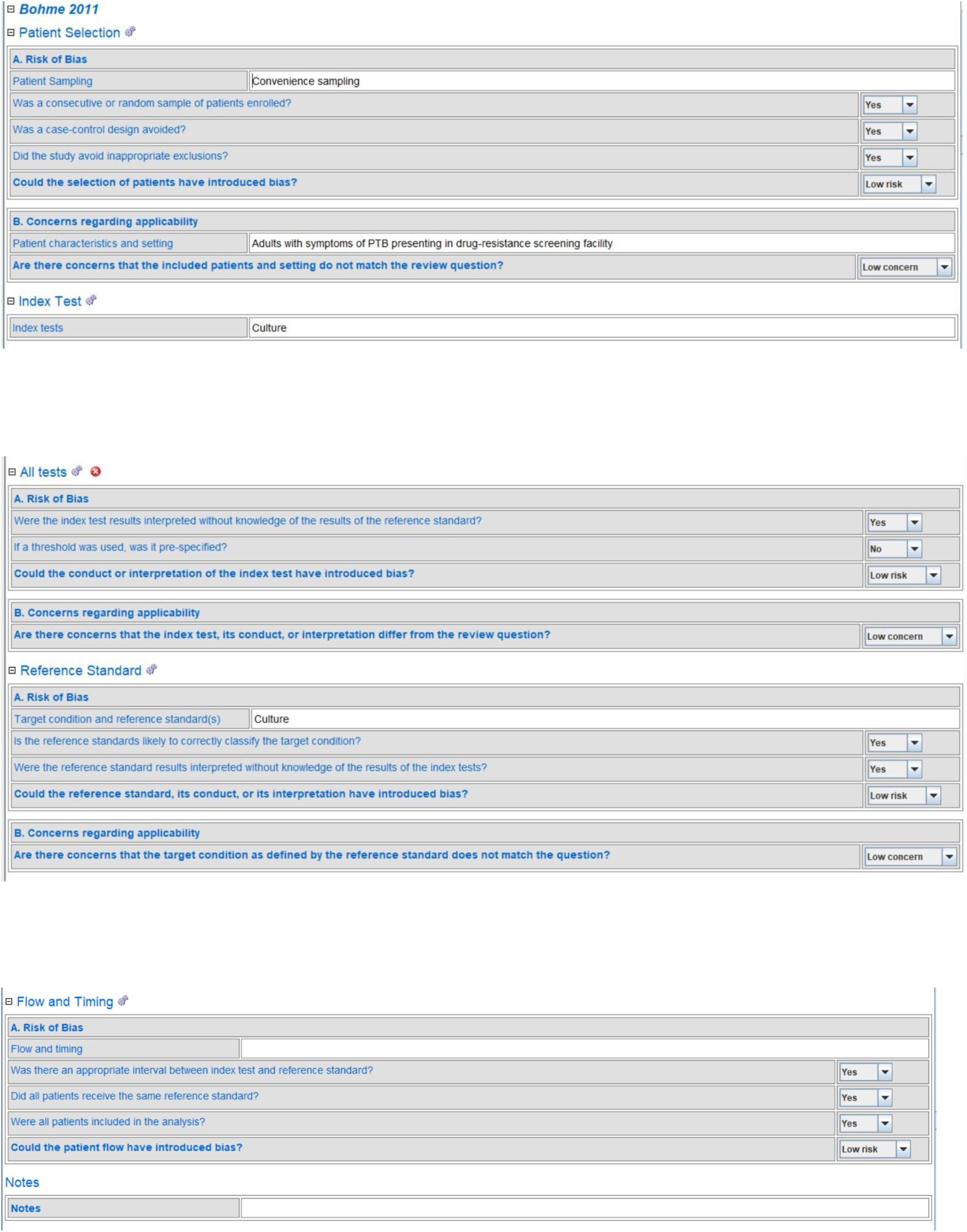

### Laserson 2005

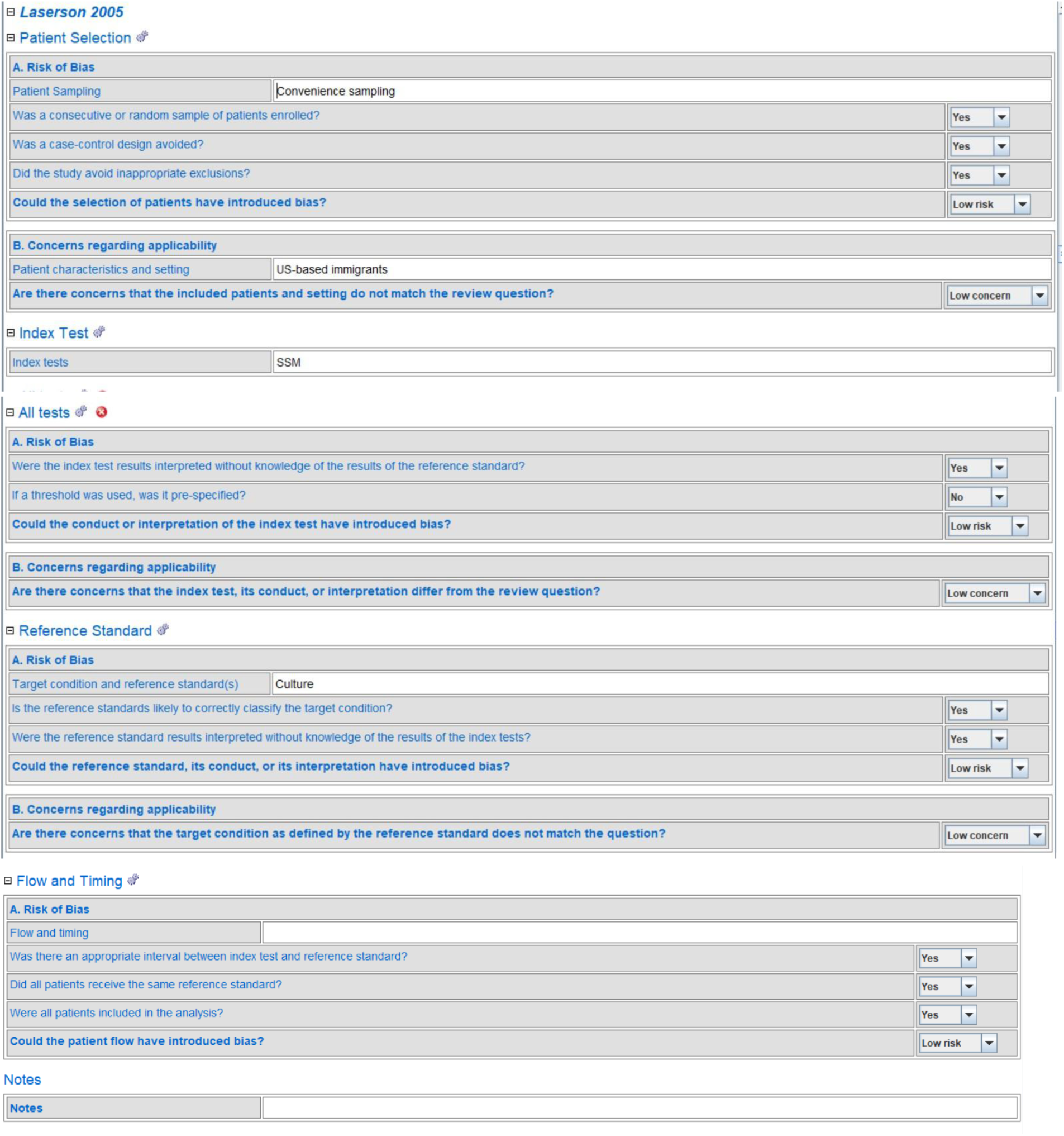

### Bellen 2003

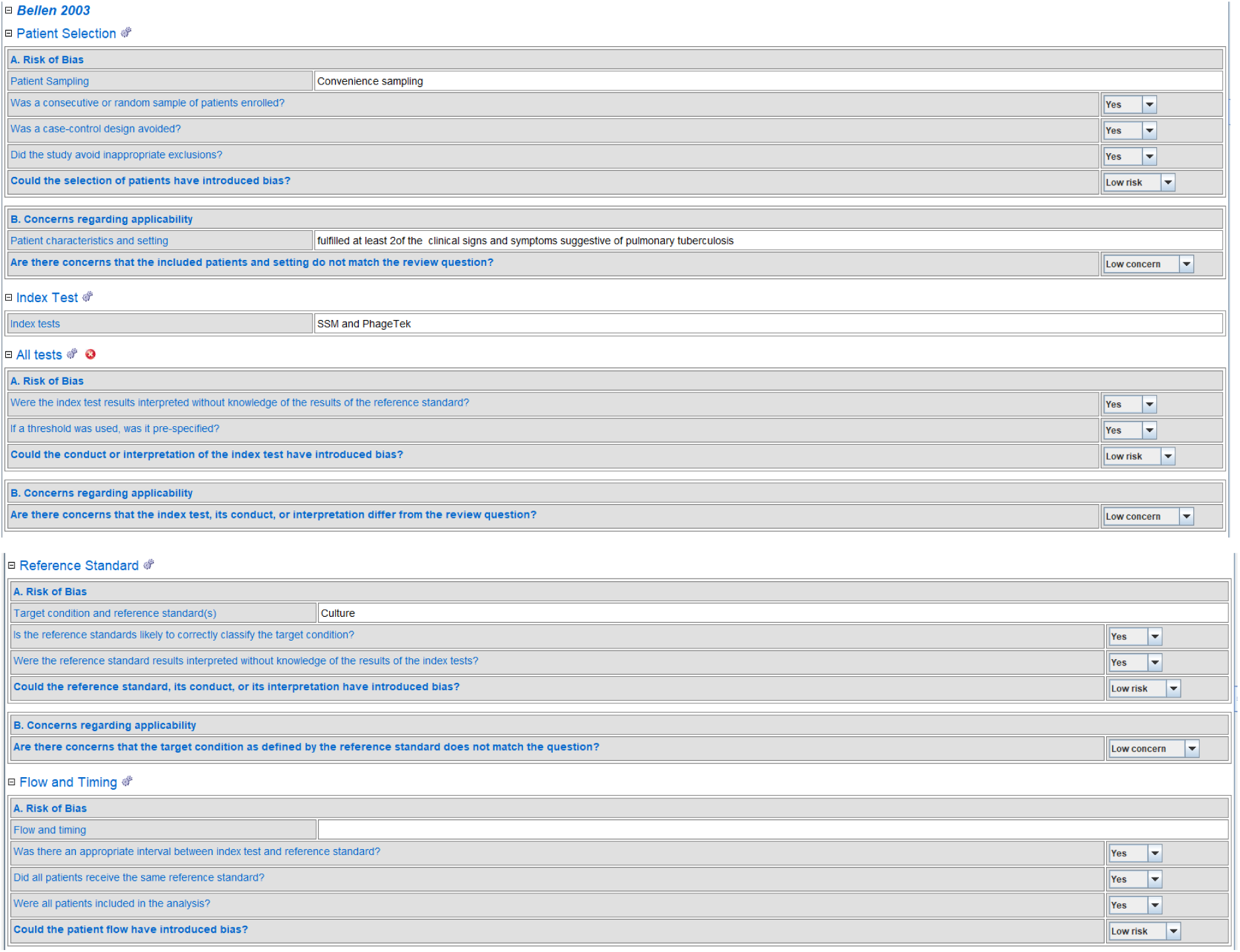

## Appendix B. MOOSE Checklist

| Reporting Criteria | Reported (Yes/No) | Reported on Page No. |
| --- | --- | --- |
| Assessment of study quality, including blinding of quality assessors; stratification or regression on possible predictors of study results | Yes |  |
| Assessment of heterogeneity | Yes |  |
| Description of statistical methods (eg, complete description of fixed or random effects models, justification of whether the chosen models account for predictors of study results, dose-response models, or cumulative meta-analysis) in sufficient detail to be replicated | Yes |  |
| Provision of appropriate tables and graphics | Yes |  |
| **Reporting of Results** |  |  |
| Table giving descriptive information for each study included | Yes |  |
| Results of sensitivity testing (eg, subgroup analysis) | Yes |  |
| Indication of statistical uncertainty of findings | Yes |  |
| **Reporting of Discussion** |  |  |
| Quantitative assessment of bias (eg, publication bias) | Yes |  |
| Justification for exclusion (eg, exclusion of non-English-language citations) | Yes |  |
| Assessment of quality of included studies | Yes |  |
| **Reporting of Conclusions** |  |  |
| Consideration of alternative explanations for observed results | Yes |  |
| Generalization of the conclusions (ie, appropriate for the data presented and within the domain of the literature review) | Yes |  |
| Guidelines for future research | Yes |  |
| Disclosure of funding source | Yes |  |

Once you have completed this checklist, please save a copy and upload it as part of your submission. DO NOT include this checklist as part of the main manuscript document. It must be uploaded as a separate file.

Retrieved from: https://www.elsevier.com/__data/promis_misc/ISSM_MOOSE_Checklist.pdf

